# Glial Maturation and Immune Landscape Dynamics in *MN1::PATZ1* Fusion-Positive CNS Tumor Recurrence

**DOI:** 10.64898/2026.02.19.26345901

**Authors:** Emon Nasajpour, Ruolun Wei, Dena Panovska, John Newman, A. Geoffrey Lyle, Ana Filipa Geraldo, Helena C.M. Oft, Yao Lulu Xing, Zhi-Ping Feng, Holly Beale, Ellen T. Kephart, Brandon Bui, Tejas Dhami, Leif Rabin, Hannes Vogel, Kelly M. Mahaney, Cynthia J. Campen, Katherine J. Ryan, Brent A. Orr, David A. Solomon, Olena Vaske, Claudia K. Petritsch

**Affiliations:** Department of Neurology and Neurological Sciences, Stanford University, Palo Alto, CA 94305, USA. [EN, DP, CKP, CJC]; Department of Neurosurgery, Stanford University School of Medicine, Palo Alto, CA 94305, USA. [EN, DP, CKP, KMM]; Division of Neuropathology, Department of Pathology, Stanford University School of Medicine, Palo Alto, CA 94305, USA; Department of Molecular, Cell and Developmental Biology, University of California, Santa Cruz, Santa Cruz, CA, USA. [AGL, HB, ETK, OV]; UC Santa Cruz Genomics Institute, Santa Cruz, CA, USA. [AGL, HB, ETK, OV]; Department of Radiology, Stanford University School of Medicine, Palo Alto, CA 94305, USA; Department of Neurology, University of Pittsburgh, 3501 Fifth Avenue, Pittsburgh, PA, 15123, USA. [HCMO]; Pittsburgh Institute for Neurodegenerative Disorders, University of Pittsburgh, Pittsburgh, PA, 15123, USA. [HCMO]; The Australian National University Bioinformatics Consultancy, John Curtin School of Medical Research, The Australian National University, ACT 2600, AU; Stanford University, Stanford, CA, 94305, USA; Maternal & Child Health Research Institute, Stanford University School of Medicine, Stanford, CA, 94305, USA. [KMM, CJC]; Division of Pediatric Neurology, Lucile Packard Children’s Hospital, Palo Alto, CA 94304, USA. [KJR, CJC]; Department of Pediatrics, Division of Hematology, Oncology, Stem Cell Transplantation & Regenerative Medicine, Stanford University, Palo Alto, CA 94305, USA. [KJR]; St. Jude Children’s Research Hospital, Division of Anatomic Pathology and Neuropathology, Memphis, TN 38105, USA. [BAO]; Stanford Cancer Institute, Stanford University School of Medicine, Stanford, CA94305, USA. [CKP]; Stanford Cancer Model Development Center, Stanford University School of Medicine, Stanford, CA 94305, USA. [CKP]

**Author notes:** Corresponding Author Claudia K. Petritsch, PhD, Departments of Neurosurgery, Neurology and Neurosciences, Stanford Cancer Institute, Stanford Cancer Model Development Center, Stanford University School of Medicine, Lorry I. Lokey Stem Cell Research Building, G1169, 265 Campus Drive, Stanford, CA 94305, USA. Equal contribution (Emon Nasajpour, Ruolun Wei, Dena Panovska, John Newman, A. Geoffrey Lyle).

**Keywords:** *MN1::PATZ1* fusion, longitudinal multi-omics, tumor maturation, tumor inflammation and immune activity

## Abstract

**Background:** *PATZ1* fusion-positive central nervous system (CNS) tumors frequently harbor *MN1::PATZ1* fusions as driver mutations, provisionally classified as a rare DNA methylation class of low-grade neuroepithelial tumors. Radiographically, they resemble pilocytic astrocytomas with tumor and cystic components, but their supratentorial cortex location and higher recurrence rates are distinguishing features. An intermediate clinical course, despite focal high-grade histopathology, underscores the need for longitudinal molecular and immune analyses to refine classification and standard therapy.

**Case Summary:** A female pediatric patient presented with neurological symptoms, including headache and right upper extremity weakness. MRI revealed a large cystic lesion in the left frontal lobe, leading to a differential diagnosis of low-grade glioma and ependymoma. Genomic analysis identified an *MN1::PATZ1* fusion. The tumor recurred after gross total resection prompting a second resection. Transcriptomic and histopathologic assessments identified multi-glial lineage, and high-grade features closely related to adult glioblastoma alongside pro-inflammatory activity in the primary tumor. The recurrent tumor showed reduced malignancy, and oligodendroglioma-like features. Increased MHC gene expression, immune checkpoint receptors (*PDCD1, CTLA4, TIGIT,TIM3*), T cell regulators (*CXCR6*), and elevated macrophage frequency, coupled with reduced PD-L1 in the recurrent tumor, suggest a complex anti-tumor immune response constrained by T cell dysregulation. This case, along with two other *MN1::PATZ1* fusion-positive tumors, identifies a distinct transcriptomic subtype separate from circumscribed astrocytic glioma, highlighting upregulation of growth factor receptor pathways, like PI3K/AKT, and immune dysfunction linked to recurrence.

**Conclusion:** Longitudinal multi-omics analyses of recurrent *MN1::PATZ1* fusion-positive CNS tumors revealed tumor maturation, immune dysfunction, and potential therapeutic targets.

**Introductory Paragraph:** *PATZ1* fusion-positive central nervous system (CNS) tumors are rare, predominantly pediatric and frequently recurrent neoplasms provisionally classified as neuroepithelial tumors. Their pronounced histopathological and clinical heterogeneity, along with limited immunological characterization complicates their treatment standardization. We report a new case of an *MN1::PATZ1* fusion-positive CNS tumor with recurrence, highlighting its radiographic similarities to low-to-intermediate grade pediatric glioma. Longitudinal multi-omics analyses of this case, along with additional *MN1::PATZ1* fusion-positive CNS tumors, however, delineates a transcriptome subtype resembling adult high-grade glioma, with activated oncogenic and pro-inflammatory programs. The recurrent tumor exhibits features of decreased malignancy and enhanced glial differentiation, phenotypically shifting towards oligodendroglioma, suggesting tumor maturation. This was accompanied by increased antigen presentation programs, indicating immune engagement, while increased immune checkpoint expression and microglia/macrophage frequency indicate T cell exhaustion and immunomodulation, respectively. This longitudinal study highlights potential therapeutic strategies targeting both the tumor and its immune environment in *MN1::PATZ1* fusion-positive CNS tumors.

## Introduction

Neuroepithelial central nervous system (CNS) tumors, *PATZ1* (POZ/BTB and AT hook containing zinc finger 1) fusion-positive (NET-*PATZ1*) lack WHO classification due to marked clinical and histopathologic heterogeneity. ^1–4^ Common fusions with the transcriptional activator *MN1* (Meningioma1) or *EWSR1* (Ewing Sarcoma breakpoint region 1), likely caused by chromosome 22 chromothripsis, share a characteristic DNA methylation profile. ^1,3,5^ *MN1::PATZ1* fusion-positive CNS tumors fall into two distinct transcriptional subgroups,neuroepithelial and sarcomatous, which are molecularly discrete from astroblastoma. ^6,7^

Radiographically, these tumors resemble juvenile pilocytic astrocytoma but are distinguished by their supratentorial location; their circumscribed nature facilitates gross total resection. ^1,4^ These tumors typically associate with frequent recurrences and favorable outcomes, indicating a low-to-intermediate malignancy grade, despite their focal high-grade histopathology. ^1^Standardizing therapy is challenged by clinicopathological heterogeneity, limited molecular data, and an undefined tumor immune microenvironment, particularly in recurrent disease.

### Case Report and Analyses

A female pediatric patient presented with headache, nausea, vomiting, fatigue, right upper extremity weakness, and aphasia. Magnetic resonance imaging (MRI) showed a large intraparenchymal cystic lesion in the left frontal lobe of the brain with an anterior mural/peripheral solid component without restricted diffusion, surrounded by mild vasogenic edema (Figures 1A B). Initial differential diagnoses included low-grade astrocytomas and ependymoma. The patient underwent gross total resection without adjuvant treatment (Figures 1A-B). A small nodule with progressive growth at the surgical margin suggested recurrence, leading to a second resection (Figures 1A-B; Table S1).

**Figure 1.**
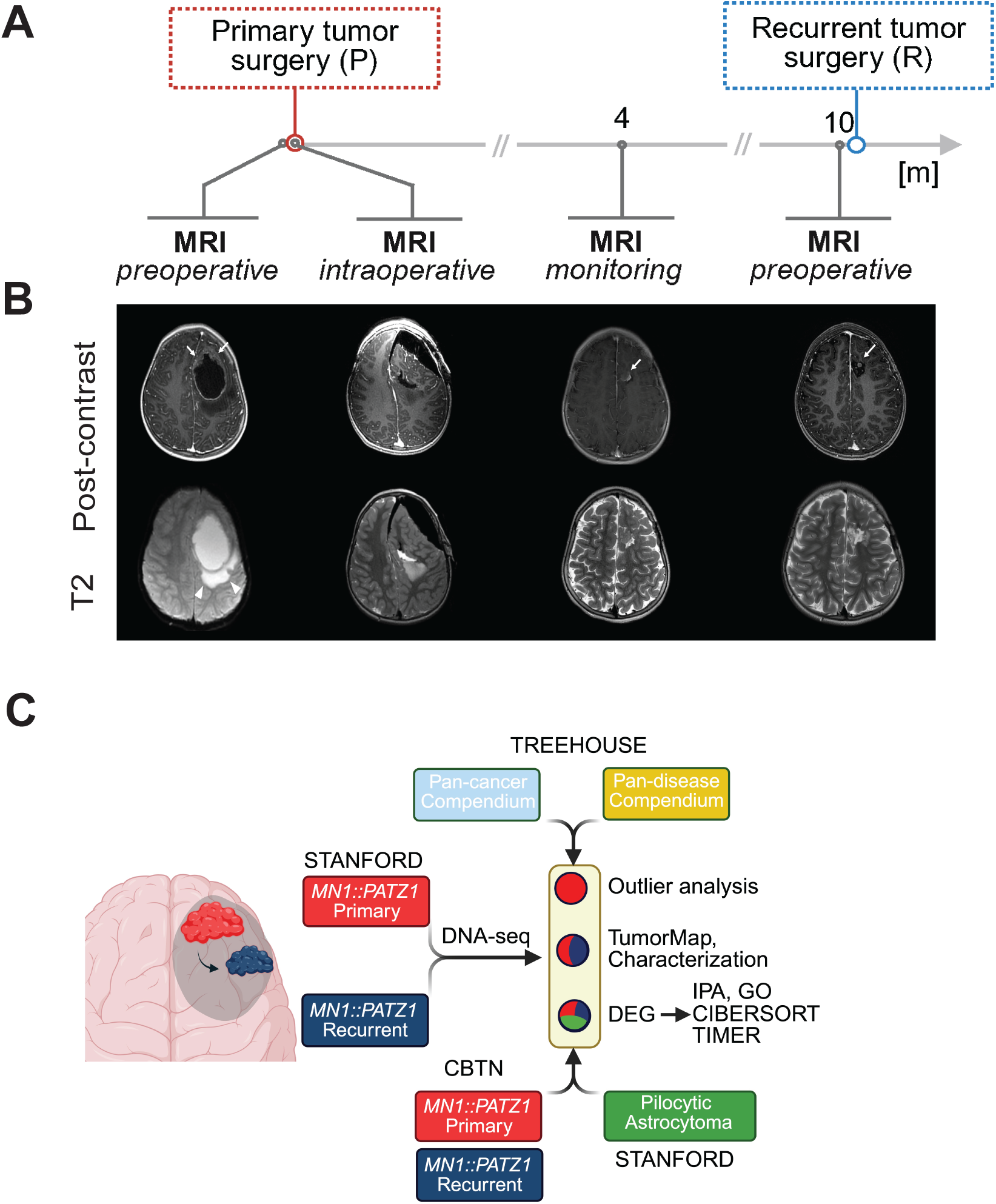
Longitudinal Clinical and Radiographical Representation of an *MN1::PATZ1* fusion-positive CNS tumor. **(A)** Clinical and surgical timeline of patient highlighting imaging, surgical intervention, and monitoring. **(B)** Longitudinal neuroimaging findings. Images include axial post-contrast T1-weighted imaging, and axial T2-weighted imaging. Preoperative MRI demonstrates a cystic lesion with an anterior solid peripheral solid component in the left frontal lobe (67 x 57 x 59 mm, AP x TR x CC) with moderate surrounding vasogenic edema (arrowheads) and right midline shift of approximately 1.7 cm. The cystic component shows peripheral enhancement while the anteriorly located small solid component (arrows) exhibits solid heterogeneous enhancement without absent diffusion restriction. Intraoperative MRI shows gross total resection of the lesion, without major complications. Monitoring/post-operative MRI obtained 4 months after surgery depicts a small solid nodule measuring 9 x 7 x 7 mm at the superomedial aspect of the surgical margin, suggesting tumor recurrence (arrow). After surgical re-intervention, a tiny, rounded area of enhancement was identified at early follow-up, remaining stable 10 months after the second procedure (arrow). **(C) Schematic detailing approaches and cases analyzed.** Figures 1A and 1C were generated in BioRender. A publication license was obtained. https://BioRender.com/tbuai2v.

#### MN1::PATZ1 fusion and unique DNA methylation profile identified by targeted DNA sequencing and methylation profiling

The patient’s family consented to donate tumor specimen under Stanford Institutional Review Board (IRB) approval. Patient-matched tissues were examined using a targeted DNA sequencing panel that detected the *MN1::PATZ1* fusion, confirmed through re-alignment and manual review. This fusion creates a chimeric oncogene, consistent with prior reports (Figures S1A-B; Table S2). ^2,4^ The DNA methylation profile did not match tumors without *PATZ1* mutations, supporting its classification as a new DNA methylation group. Copy number analyses showed no large alterations, but significant changes on chromosome 22 indicated chromothripsis, consistent with previous reports on *PATZ1*-mutant CNS tumors (Figure S1C). ^3^

#### Molecular profiling reveals outlier genes and glial differentiation in MN1::PATZ1 fusion-positive CNS tumors

Extensive molecular profiling was performed for this case (Figure 1C). Molecular transcriptome profiling to the Treehouse cancer compendium v11 polyA (n=12747) clustered the samples with the brain tumor cohort (n=971) (Figure 2A; Supplemental File; Methods) ^8^ and revealed most close correlation of the primary tumor with adult type diffuse glioma; and primitive neuroectodermal tumor (PNET) - now WHO classified as CNS embryonal tumors (Figures 2B; S2A-C). Comparing the primary *MN1::PATZ1* fusion-positive tumor transcriptome to *1)* the whole Treehouse reference compendium (pan-cancer) and *2)* the Treehouse brain tumor-focused cohort (pan-disease), identified 214 pan-cancer outlier genes (189 significantly upregulated; 25 significantly downregulated) and 147 pan-disease outlier genes (79 significantly upregulated; 68 significantly downregulated) (Table S3). Key pan-disease outliers included previously reported upregulated transcripts *IGF2*, *GATA2*, *PAX2*, *CSPG4,* ^2^ new transcripts for *CDC34*, *PATZ1*, and oligodendroglia lineage (*OLIG1/2, SOX10*), differentiation (*PLP1, CNP*), and astrocyte maturation (*AQP4, GFAP*) transcripts (Figure 2C, Figures S2D-M), indicating tumor glial differentiation. Expression outlier genes are not provided for the recurrent tumor sample due to insufficient number of mapped unique exonic non-duplicate reads, which did not pass strict Treehouse *Tumor Mapping and Comparative Analysis of RNA expression (CARE)* quality control metrics (Supplemental File; Methods). Gene ontology (GO) analysis revealed enrichment in synaptic signaling, neural progenitor proliferation, and astrocyte projections, consistent with high synaptophysin and GFAP positivity by IHC (Figures S3A-B).

**Figure 2.**
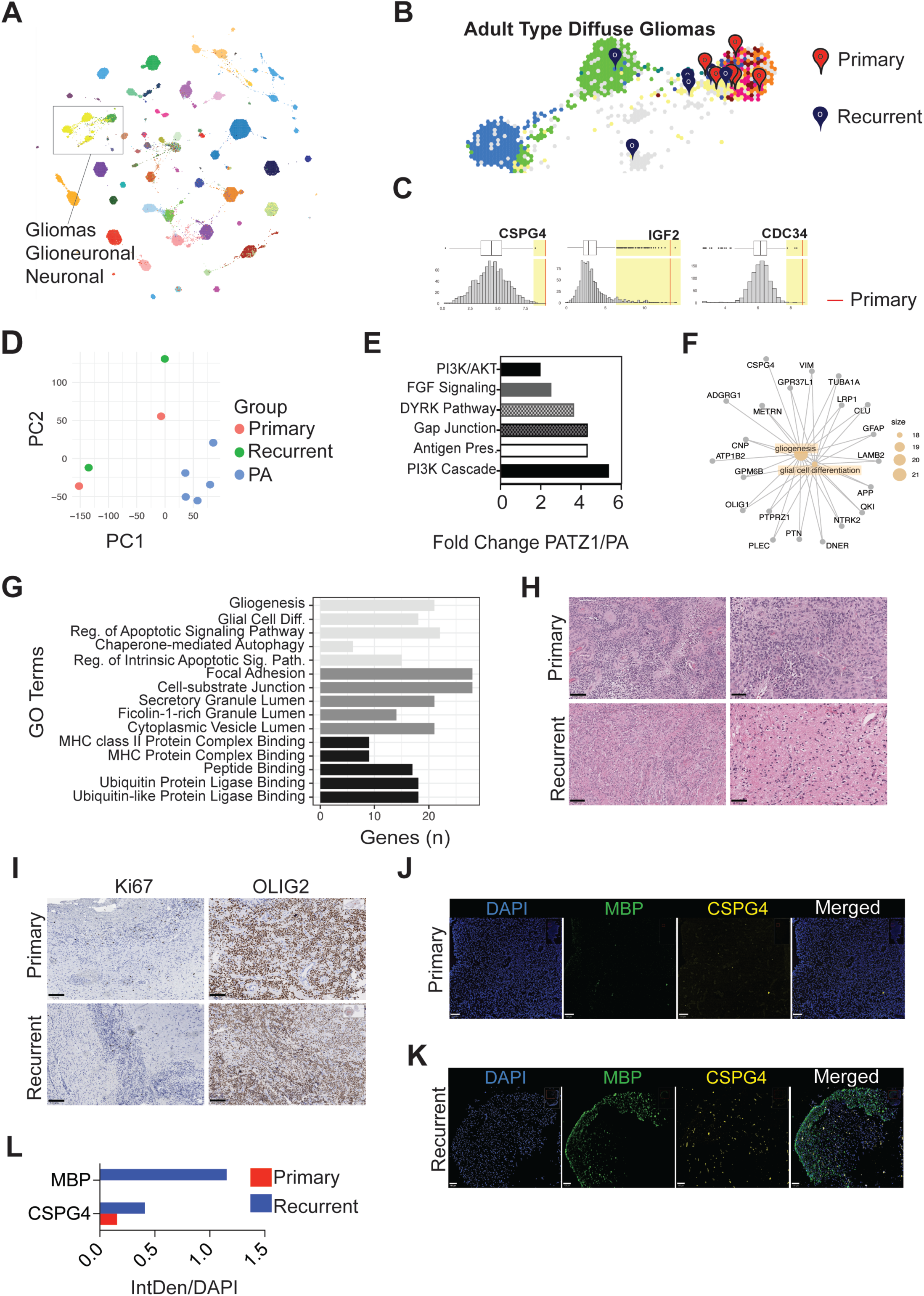
Transcriptomic and Histopathologic Analyses Reveal Adult-Type Glioma Similarity with Upregulated Cancer and Cell Cycle/Proliferation Pathways. **(A)** Treehouse v11 polyA cancer compendium containing 12,747 pediatric and adult tumor cases. Tumors are grouped based on RNA-seq similarities. The rectangle highlights enlarged gliomas (yellow) and glioblastoma, *IDH*-wildtype (GBM) (green). **(B)** Transcriptomic neighbor analysis of both the primary and recurrent *MN1-PATZ1* fusion-positive CNS tumor. Zoomed-in image of the glioma and glioblastoma module representing adult type diffuse gliomas, including glioblastoma, *IDH*-wildtype (yellow), encompassing mesenchymal (orange), neural (red), and proneural (purple) subtypes, astrocytoma, *IDH-*mutant without codeletion (green), and oligodendroglioma, *IDH*-mutant with 1p/19q-codeletion (blue). Each colored dot represents an individual patient’s tumor RNA-seq data. The primary tumor sample (red pin) has nearest transcriptomic neighbors of glioblastoma (GBM), astrocytoma grade 3 (G3), and PNET, while the recurrent tumor sample (blue pin) has nearest transcriptomic neighbors of oligodendrogliomas and astrocytomas, grades 2 and 3 (G2/G3), and oligoastrocytoma grade 3 (G3). The correlation index was lower in the recurrent sample (0.86-0.89) in comparison to the primary (0.88 - 0.91). The terms “PNET” and “oligoastrocytoma” are no longer used in the WHO CNS5 (2021) tumor classification system. **(C)** Representative histograms of the outliers expression including *CSPG4*, *IGF2* and *CDC34* in the primary *MN1::PATZ1* fusion-positive CNS tumor (STN126), as compared to the v11 polyA brain tumor cohort (n = 971) samples. **(D)** Principal component analysis (PCA) plot comparing four MN1::PATZ1 fusion-positive CNS tumors (including primary and recurrent patient-matched pairs, plus two additional cases from the Treehouse public dataset) with five pilocytic astrocytomas samples derived from our in-house cohort. **E)** Selected upregulated canonical pathways in *MN1::PATZ1* fusion-positive CNS tumors (n=4; STN10126, STN10156, THR60 5286, THR60 5285) versus pilocytic astrocytomas (PA, n=5), as determined by Ingenuity Pathway Analysis (IPA) of the top differentially expressed genes. See Supplemental Table S1 for patient details. **(F)** Cnetplot cross-validation of genes in “gliogenesis” and “glial cell differentiation” GO terms in the recurrent *MN1::PATZ1* fusion-positive CNS tumors. **(G)** GO pathway enrichment analysis of upregulated genes in the recurrent *MN1::PATZ1* fusion-positive CNS tumors. Antigen presentation is enhanced in the recurrent *MN1::PATZ1* fusion-positive tumors, alongside glial differentiation, and MHC and MHC class II expression. **(H)** Histopathologic analyses of the primary (STN10126) and recurrent (STN10156) *MN1::PATZ1* fusion-positive CNS tumor specimen using H&E staining. Low-power image showing hypercellular brain parenchyma with neoplastic proliferation of glial cells with perivascular anuclear zones in the primary tumor. High-power images showing perivascular pseudorosettes, and areas with more monomorphic, round to ovoid cytology. High grade features including microvascular proliferation and abundant palisading necrosis were present in the primary tumor. The recurrent tumor was nearly entirely composed of neoplastic glial cells with more uniform round to ovoid nuclei arranged in perivascular pseudorosettes and astroblastomatous rosettes. Necrosis and microvascular proliferation were absent in the recurrent tumor specimen. Scale bars low magnification = 100 μM; Scale bars high magnification = 50 μM. **(I)** Immunohistochemistry for Ki67 of the primary and recurrent *MN1::PATZ1* fusion-positive CNS tumor showing a proliferation index of approximately 10% in the primary tumor and about 2% in the recurrent tumor. Immunohistochemistry for OLIG2, which marks oligodendroglial lineage cells of the primary and recurrent *MN1::PATZ1* fusion-positive CNS tumor. OLIG2 highlights approximately 70% of the cells in the primary tumor, especially the cells with oligodendroglial-like morphology arranged in ependymoma-like perivascular pseudorosettes. OLIG2 highlights approximately 80% of cells in the recurrent tumor. Scale bars = 100 μM. **(J, K)** Representative immunofluorescence images of the primary **(J)** and recurrent **(K)** *MN1::PATZ1* fusion-positive tumors stained with oligodendrocyte progenitor and differentiation specific antibodies, including the oligodendrocyte progenitor marker CSPG4/NG2 and the myelinating oligodendrocyte marker myelin basic protein MBP. Scale bars = 400 μM. (**L**) Quantification of (J). Barplot of MBP and CSPG4 intensity normalized to DAPI-positive nuclei in primary and recurrent sample. The experiment was performed in two technical replicates (n=2).

#### MN1::PATZ1 tumors are molecularly distinct from pilocytic astrocytomas, exhibiting activated oncogenic pathways linked to recurrence

To investigate further molecular distinctions from pediatric low-grade gliomas (pLGG), we performed principal component analysis (PCA) on transcriptomes from *MN1::PATZ1* (n = 4) and pilocytic astrocytomas (PA) (n = 5), which showed no clustering (Figure 2D, Table S1). Ingenuity Pathway Analysis (IPA) on *MN1::PATZ1* versus PA transcriptomes revealed heightened activation of PI3K–AKT signaling, further confirmed by KEGG (Figures 2E, S3C), and other pathways, including antigen presentation, suggesting that these features may contribute to their recurrence potential (Figure 2E; Table S4).

Given the rarity of reported recurrences and lack of documented molecular differences between patient-matched primary and recurrent tumors, we conducted comparative mapping of the paired *MN1::PATZ1* fusion-positive tumors. The recurrent tumor showed highest similarity to oligodendroglioma, astrocytoma, and oligoastrocytoma—an entity that is no longer used by the 2021 WHO CNS tumor classification and was replaced by oligodendroglioma and astrocytoma with assigned grades 2-3 (Figure 2B, Figures S2A-C). Differential gene expression and GO analyses revealed enrichment for gliogenesis and glial differentiation in recurrent tumor (Figure 2F-G).

Histopathologically, the primary tumor demonstrated high-grade glioma features resembling glioblastoma, including irregular hyperchromatic nuclei, dense fibrillary processes, both ependymoma-like perivascular pseudorosettes and astroblastomatous rosettes, several foci of palisading necrosis, and well-developed microvascular proliferation. The recurrent tumor exhibited a distinct morphology with oligodendroglial characteristics, retaining perivascular pseudorosettes and astroblastomatous rosettes, but lacking high-grade features Figure 2H). Immunohistochemistry (IHC) showed low but focal Ki67 proliferation rates reaching about 10% in the primary and 2% in the recurrent tumor (Figure 2I), with a predominant glial lineage indicated by markers glial fibrillary acidic protein (GFAP), vimentin, and OLIG2 (70-80%), with minimal synaptophysin and no intercellular reticulin or desmin (Figures 2I, S3A), indicating a predominantly glial lineage tumor lacking overt neuronal differentiation. Increased expression of mature oligodendrocyte marker myelin basic protein (MBP) and GFAP in the recurrent tumor confirmed enhanced glial differentiation, corroborating the RNA seq findings (Figures 2F, 2J-L) (Figure S3A). Increased expression of oligodendroglial progenitor cell marker (CSPG4) in combination with decreased OLIG2 indicates enhanced astrocytic cell states. These findings indicate a multi-glial lineage tumor with robust glial specification and enhanced glial differentiation in the recurrent lesion, suggesting a shift towards a more mature, less malignant phenotype.

#### Inflammatory signatures and T cell suppression as potential drivers for tumor recurrence

We characterized the tumor immune microenvironment (TIME) in *MN1::PATZ1* fusion-positive CNS tumors by IPA, revealing increased pro-inflammatory cytokines (*IL6, IL11, CXCL8/IL8*) in primary but not in recurrent tumor (Figure 3A; Supplemental Table S7) and heightened antigen presentation programs in *MN1::PATZ1* fusion-positive tumors compared with PAs (Figure 2E) and further elevated in the recurrent tumor, alongside other immune pathways, as shown by GO analyses (Figure 2G) and confirmed by a Cnetplot (Figure 3B). Immune checkpoint regulator transcripts (*PDCD1*, *CTLA4, TIGIT,* and *TIM3*) and the T cell recruiting chemokine *CXCR6*, were elevated in recurrence, suggesting heightened T cell infiltration alongside immune suppression as a response to inflammation (Figure 3C). Tumor Immune Estimation Resolution (TIMER) analysis showed increased macrophages, dendritic cells, and B cells, with slight CD8+ T cell elevation, and decreased CD4+ T cells and neutrophils in the recurrent tumor (Figure 3D) corroborated by CIBERSORT deconvolution (Figure S4A) and validated by IHC confirming increase in CD8+ (1% in the primary; 2-3% in the recurrent tumor) and CD68+ (approximately 15% to 25%) cells (Figure 3E, S4A) and minimal CD4+ cell presence (Figure S4B). The *programmed cell death protein 1* (*PDCD1*) transcript, encoding the immune-inhibitory PD-1 receptor, was highly expressed (Figure 3C) and identified as an upregulated outlier by CARE (Figure 3G). Both the PD-1 ligand PD-L1 transcript and protein were also highly expressed in the primary tumor (Figures 3E-F), suggesting potential responsiveness to therapeutic PD-1 checkpoint blockade. To explore this further, we interrogated the transcriptomes of *MN1::PATZ1* fusion-positive CNS tumors (n=4) and PAs (n=5) for an IFN--related mRNA profile known to predict clinical response to PD-1 blockade. ^9^ This tumor inflammation signature integrates 19 genes comprising several aspects of the immune response in a single numerical value. We found non-uniform upregulation of six signature transcripts in the *MN1::PATZ1* fusion-positive tumors compared to more uniform upregulation of 11-15 transcripts in the PAs. Interestingly, heatmap analysis indicated distinct immune profiles based on *MN1::PATZ1* fusion status (Figure 3F). Relative to PAs, *MN1::PATZ1* fusion-positive tumors exhibited non-uniform upregulation of transcripts, including for inhibitory immune checkpoints *PDCD1, TIGIT,* immunosuppressive markers *IDO1,* and chemokine receptor *CXCR6*, accompanied by downregulation of transcripts for MHC class II mediated antigen presentation (*HLA-DRB1, HLA-DQA*), inhibitory ligands, co stimulatory/co inhibitory receptors (*CD27*), and T cell activation markers (*CD8, STAT1*).

**Figure 3.**
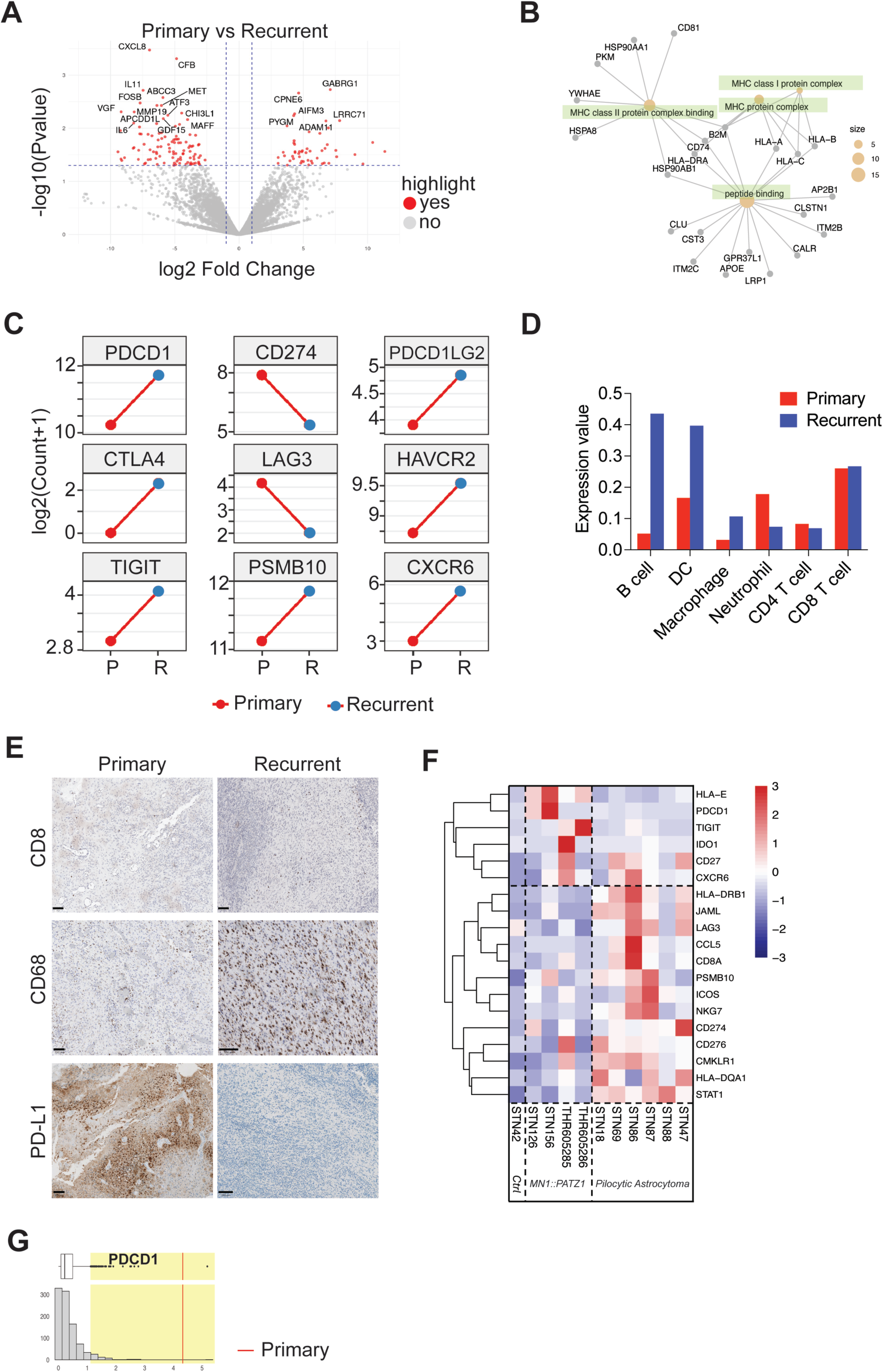
Dynamic Changes in the Tumor Immune Landscape of *MN1::PATZ1* Fusion-positive CNS Tumors. **(A)** Volcano plot visualizing relative differential gene expression comparing the transcriptomes of primary (n=2; STN10126, THR60 5286) with recurrent (n=2; STN10156, THR60 5285) *MN1::PATZ1* fusion-positive CNS tumors. The x-axis shows the log2 of the Fold Change in expression levels between primary (left) and recurrent (right). The y-axis shows the negative logarithm base 10 (Log10) of the p-value associated with each transcript. The graph depicts unadjusted, raw p-values for an n=2 per condition, FDR=1. Red highlighted transcripts are significantly upregulated in primary or recurrent tumors. **(B)** Cnetplot cross-validation of genes in immune-related GO pathways in the recurrent *MN1::PATZ1* fusion-positive CNS tumor (n=1; STN10156). **(C)** Immune modulatory transcripts trends in the patient-matched primary (n=1; STN10126) and the recurrent (n=1; STN10156) *MN1::PATZ1* fusion-positive CNS tumor. The transcripts of *PDCD1, CTLA4, TIGIT, PSMB10, PDCD1LG2, HAVCR2/TIM3,* and *CXCR6* are upregulated in the recurrent tumors, while others (*CD274/PD-L1* and *LAG3*) are downregulated with recurrence. **(D)** TIMER deconvolution of the patient-matched primary (n=1; STN10126) and the recurrent (n=1; STN10156) *MN1::PATZ1* fusion-positive CNS tumor, suggesting shifts in immune-cell composition. **(E)** Immunohistochemical staining for CD8 (top panels), CD68 (middle panels), and PD-L1 (bottom panels) was performed on primary and recurrent *MN1::PATZ1* fusion-positive CNS tumors. The staining results reveal intratumoral CD8+ T-cells in both the primary tumor (<1%) and the recurrent tumor cores, indicative of immune altered, immune compromised tumors (2-3%). The macrophage marker CD68 is largely negative in the primary tumor, whereas robust macrophage infiltrates are evident in the recurrent tumor. Furthermore, the PD-L1 tumor proportion score (TPS) in the primary tumor is approximately 5% (considered positive, as it is >1% but <50%), while the recurrent tumor shows a TPS <1%, indicating no significant PD-L1 staining. Scale bars = 100 μM. **(F)** Heatmap comparing immune-checkpoint expression in *MN1::PATZ1* fusion-positive tumors (n=4; STN10126, STN10156, THR60 5285 S01, THR60 5286 S01) versus a control cohort, consisting of pilocytic astrocytomas (n=6; STN10018, STN10069, STN10086, STN10087, STN10088, STN10047), and normal control from non-malignant brain tissue of a craniotomy to remove epileptic foci (n=1; STN10042). Non-uniform upregulation of immune-inhibitory receptors (*PD-1, TIGIT*), the tryptophan-catabolizing enzyme *IDO1*, the immunoproteasome subunit *PSMB10*, and the chemokine receptor *CXCR6,* is contrasted by downregulation of genes central to MHC class II–mediated antigen presentation (*HLA-DRB1, HLA-DQA1*), inhibitory ligands (*PD-L1, B7-H3*), co-stimulatory/co-inhibitory receptors (*ICOS, LAG3*), and effector markers (*CD8A, NKG7, CCL5, CMKLR1, JAML*) in *MN1::PATZ1* fusion-positive tumors. **(G)** Analysis showing that *PDCD1* is an upregulated outlier transcript in primary (red line) *MN1::PATZ1* fusion-positive tumor (STN10126) in comparison to a pan-disease panel. Note, that the recurrent sample did not pass the QC threshold.

This suggests an anti_tumor immune response balanced with T_cell exhaustion, and an attenuated anti-tumor immune-response due to macrophage recruitment in *MN1::PATZ1* fusion positive tumors compared to PAs (Figure 3F).

## Discussion

There is no established standard of care for *MN1::PATZ1* fusion-positive CNS tumors beyond surgical intervention, with management typically individualized within a multidisciplinary setting. Focal radiation may be considered for unresectable localized high-grade cases, while systemic chemotherapy plays a limited role due to the tumor’s indolent biology and low proliferative indices (Ki-67 often 1–5%), showing minimal clinical response. ^3^ Our data show upregulation of PI3K/AKT signaling in *MN1::PATZ1* fusion_positive tumors versus PAs, suggesting this pathway may contribute to recurrence and nominating the PI3K/AKT/mTOR axis for preclinical evaluation of pathway inhibitors, some of which are under investigation in pediatric settings. ^10^

In the new case reported here, the *MN1::PATZ1* fusion is the sole alteration of known significance in these very rare tumors, akin to other tumors driven by fusion oncoproteins, ^2^ suggesting that this fusion may also generate tumor antigenicity (Supplemental Table S1, Figure 2E). *MN1* overexpression with varying tumor grade is a promising predictor of survival of glioma patients. ^11^ Our longitudinal multi-omics analyses of a paired *MN1::PATZ1* fusionpositive CNS tumor provide the first reported case with molecular analyses, highlighting changes in tumor states, types, and malignancy grades with recurrence. The enrichment of glial differentiation markers, including MBP and GFAP, and the loss of malignant features indicate that the recurrent tumor is more mature, acquiring glial differentiation traits. Notably, this maturation occurs in the absence of any adjuvant treatment and may explain the relatively indolent course in some patients despite frequent local recurrences, considering a median progression-free survival of 91 months. ^1^ Our PCA data and pathway analyses reinforce the distinct separation from PAs, and tumor mapping placed our case primarily within adult-type gliomas, despite being pediatric. These findings corroborate that *MN1::PATZ1* fusion-positive CNS tumors represent a Regarding experimental therapies, Alhalabi *et al*. reported drugscreening results that identified several candidate agents, including the BCL-2/BCL-XL/BCL-w inhibitor navitoclax. ^1^Navitoclax has been shown to exhibit limited blood–brain barrier penetration in non-human primates and is associated with dose-limiting thrombocytopenia in both pediatric and adult trials, factors that substantially constrain its clinical applicability. These observations suggest that the development and preclinical evaluation of more selective BCL-xL inhibitors, potentially in combination with other approaches, remain areas for future investigation, particularly in the context of *MN1::PATZ1* fusion–positive tumors, which predominantly occur in pediatric patients.

Clinical success for immune checkpoint blockade in pediatric patients has thus far been limited to pediatric patients with mismatch repair deficient (dMMR) high-grade gliomas (HGG). We previously showed that PD-1 and/or CTLA-4 inhibitors can improve efficacy in BRAFV^600E^ mutant HGG, particularly when combined with molecular targeted therapies. ^12,13^ The tumor immune microenvironment in pediatric low-grade gliomas (pLGG) and some subgroups of pediatric HGG (pHGG), including the BRAFV600E-mutated HGG, is characterized by T cell infiltration and PD-L1 expression, which may predict a clinical response to PD-1 blockade.^12,14^ IFN-γ related mRNA signatures, Immune Pediatric Signature Score (IPASS), and tumor inflammatory signatures (TIS) have been developed as scoring systems to characterize the immune landscape of cancers, including pediatric brain cancers and are superior to tumor mutational burden and PD-L1 expression at predicting the potential effectiveness of PD1 immune checkpoint inhibition in pediatric dMMR HGG.^9,15^, Levine *et al.* found high immune activation in pediatric gliomas, particularly in pLGG versus pHGG and circumscribed over diffuse LGGs, ^14^ while our study shows that *MN1::PATZ1* fusion-positive tumors also exhibit an active immune response, though lower than PAs. The presence of T_cell infiltration and elevated pro_inflammatory cytokines (IL_6, IL_8) in the *MN1::PATZ1* primary tumor and the immune profile shifts marked by enhanced antigen presentation despite overall an immune suppression signature in the recurrent tumor, indicates an active and dynamic immune infiltrate and provides rationale to explore PD1 pathway blockade as a therapy.

Tools like CIBERSORT, TIMER have been developed to computationally assess immune cell infiltrates. We used these methods to identify the presence of cytotoxic cells (TILs, NK cells, neutrophils), although some divergence exists among them due to differences in cell annotation methods.

Our findings highlight the potential for personalized treatment strategies leveraging immune checkpoint inhibitors, while emphasizing the need for future analyses of additional patientmatched primary and recurrent tumors and validating this observation utilizing (spatial) single cell RNA sequencing tools.

## Data Availability

All data produced in the present study are available upon reasonable request to the authors

## Funding

BRAF LGG consortium research fund (C.K.P.), National Institute of Health [17X074] (C.K.P.) Bio-X Seed Grant (C.K.P), Stanford Cancer Institute (C.K.P), National Institute of Health U54 [CA 261717] (C.K.P, H.V.).

## Conflict of Interest

The authors declare no conflicts of interest in relation to this study.

## Authorship

Generated Figures 2 and 3 histopathologic slides, wrote parts of the results and figure legends, provided neuropathologic consult: EN, DS, HV, JN

Obtained reports and details about targeted sequencing data, analyzed targeted sequencing for fusion breakpoints: DS

Re-analyzed DNA sequencing data, generated Supplemental Figure 1: YLX, ZPF

Organized data analyses for Figure 2, arranged collaboration with UCSF team, isolated and submitted RNA for sequencing, advised on Figure design, wrote results for figure 2 and supplemental figure 2, provided scientific guidance, generated high resolution figures for submission: DP

Created IF data: BB

Generated DNA methylation data: BO

Created data for Figure 3 CIBERSORT and pathway analyses, wrote results, revised and edited manuscript: RW

Created DEG, Ingenuity Pathway Analyses and Volcano plot data: HCMO

Consulted in analysis of DNA sequencing data, provided methods support, assisted with Supplemental Figure 1: ZPF

Organized materials for molecular analyses and patient data: EN, TD

Gathered and interpreted MRI, reviewed manuscript: EN, TD, RW, APG

Provided surgical tissue and patient data: KM

Provided patient clinical history and input: EN, CJC, KJR

Downloaded and analyzed data from PedBioPortal: AGL

Performed tumor mapping and comparative analysis of RNA expression (outlier): HB, AGL, ETK

Conceptualized tumor mapping and comparative analysis of RNA expression (outlier), supervised bioinformatic approaches, edited drafts, and final manuscript: OMV Conceptualization, organized, and supervised data acquisition, created parts of Figures, inquired molecular analyses, finalized Figures, edited drafts and final manuscript, reviewed first and revised submission manuscript: EN, CKP

## Ethics approval

The authors confirm that written consent for submission and publication of this case report, including the images and associated text, have been obtained from the patient(s) in line with COPE guidance.

## Data availability

RNA sequencing data from this case were deposited in GEO and will be made available at manuscript acceptance. The data are available to the editors and reviewers upon manuscript submission.

## Acknowledgements

We thank Pauline Chu for image scanning, Caden Che for initial analyses of gene expression data, and Dr. Keith Ligon and Jaquelyn Jones (DFCI) for patient information. Figures 1A and C were created in BioRender. Petritsch, C. (2025) https://BioRender.com/tbuai2v (https://biorender.com/tbuai2v).

## Notes

### Competing Interest Statement

The authors have declared no competing interest.

### Funding Statement

National Institute of Health 17X074 and CA 261717

### Author Declarations

Ethics committee/IRB12625 of Stanford University School of Medicine gave ethical approval for this work.

