## Supplementary material for "Glial Maturation and Immune Landscape Dynamics in *MN1::PATZ1* Fusion-Positive CNS Tumor Recurrence": supp data

**Supplemental Figures and Tables**

**Supplemental Figure S1: Identification and validation of the *MN1::PATZ1* fusion.**

**Supplemental to Figure 1.**

UCSF 500 targeted sequencing analysis and alignment to human reference GRCh37/hg19 revealed the presence of the *MN1::PATZ1* fusion gene, which was validated by re-alignment to a different reference genome.

**(A)** Schematic diagram of the *MN1::PATZ1* fusion event based on re-analysis of targeted exome sequencing (UCSF500 Cancer Gene Panel) of the patient’s tumor tissue. Data were aligned to the human reference genome GRCh38/hg38, showing breakpoints in the intron 1 of *MN1* (chr22: 27,769,473) and exon 1 of *PATZ1* (chr22: 31,344,580). The original analysis using GRCh37/hg19 reference genome showed similar results with slightly different breakpoint coordinates (*MN1*: chr22: 28,165,461; *PATZ1*: chr22: 31,740,566). The fusion joins the N-terminal transactivation domain of *MN1* to the DNA-binding Zinc finger motifs of *PATZ1*, replacing its N-terminal PTB/BOZ protein interaction domain and AT-hook DNA-binding motif, and creating a chimeric oncogene. Fusion junctions in intron 1 of *MN1* and exon 1 of *PATZ1* caused an in-frame linkage of exon 1 of *MN1* (codons 1-1260) together with exons 1-5 of *PATZ1* (codons 342-687).

**(B)** Integrated Genomics Viewer (IGV) screenshot showing read alignments in split-screen view of the identified *MN1::PATZ1* fusion. Reads are colored by insert size (red and green reads) and pair orientation (pink and blue reads were aligned to the forward and reverse strand, respectively). The reads mapped in intron 1 of *MN1* gene paired with those mapped in exon 1 of *PATZ1* gene, with breakpoints at Chr22: 27,769,473 and Chr22:31,344,580, respectively. The read coverage is X for *MN1* and Z for *PATZ1* of the *MN1::PATZ1* fusion.

**(C)** Copy Number Variation Profile from Methylation Profiling for the primary *MN1::PATZ1* fusion-positive tumor showing a flat chromosome with no major gains or losses, except for chromosome 22. Depiction of chromosome 1 to 22. Gains/amplifications would represent positive, losses/negative deviations from the baseline in blue. 29 brain tumor regions are highlighted for easier assessment. Methylation profiling suggest that no major gains or losses occurred in this *MN1::PATZ1* fusion -positive tumor except for chromosome 22, which showed some losses.

**Supplemental Figure S2:** **Comparative Analysis of RNA Expression (CARE) and Ingenuity Pathway Analyses (IPA) Identify Known Outliers and Significantly Expressed Pathways in *MN1::PATZ1* fusion-positive CNS tumors.**

**(A, B)** Brain tumor maps colored by sample correlation index of primary **(A)** and recurrent **(B)** tumor.

**(C)** Brain tumor map colored by tumor grade: grade 2 (orange), grade 3 (green), grade 4 (blue).

**(D-M)** A comparison of the primary transcriptome and Treehouse v11 polyA pan cancer compendium (12,474 cases) as a background cohort using CARE. The gene expression level in the sample is denoted with a vertical line in the primary (red line) tumor plotted with respect to the distribution of the gene expression across the comparator cohort whereas each dot represents an individual tumor sample. The sample of the recurrent tumor did not pass the stringent UCSC quality control metrics was therefore excluded from this analysis. The x-axis is the gene expression in log2(TPM+1), and the y-axis is the number of samples in the background cohort that fit into that expression bin. The box plot is a Tukey boxplot show the interquartile range (Q1 - Q3), the median, and the whiskers are the interquartile range * 1.5.  The dots after the whisker are outliers.

CARE shows known upregulation of *PAX2* **(D)**, *IGF2* **(E)**, *GATA2* **(F)**, *PATZ1* **(G)**, *SOX10* **(H)**, *OLIG2* **(I),** *PLP1* **(J),** *CNP* **(K),** *AQP4* **(L)***, GFAP* **(M)**, (and above the 95^th^ percentile threshold for the sample).

**Supplemental Figure S3: Immunohistochemical Analyses of Primary and Recurrent *MN1::PATZ1* fusion-positive CNS tumors and Gene Enrichment analyses.**

**Supplemental to Figure 2.**

**(A) Representative immunohistochemistry staining.** GFAP and Vimentin staining was detected in both primary and recurrence. Synaptophysin stain positivity increased from 1% in the primary to 20% positivity in recurrence. Reticulin stain in primary highlights vascular basement membranes, negative for intercellular tumoral deposition. Reticulin stain in recurrence highlights vascular basement membranes, negative for intercellular tumoral deposition. Desmin staining is negative in both primary and recurrence. Scale bars = 50 μM.

**(B) Pathway analyses of outliers.** The Manhattan plot was generated using the pan-cancer outlier genes g:OST function of g:Profiler and found over-representation of genes among Gene Ontology terms, molecular function, biological pathways, and cellular components. The highly represented pathways include synapse, nervous system development, synaptic signaling and protein binding.

**(C)** Venn Diagram comparing *MN1::PATZ1* fusion-positive tumors (n=4; STN10126, STN10156, THR60 5285 S01, THR60 5286 S01) versus a control cohort, consisting of pilocytic astrocytoma (n=5; STN10018, STN10069, STN10086, STN10087, STN10088). The DEGs were analyzed by KEGG Pathway Enrichment. Unique DEGs from the primary *MN1::PATZ1* fusion-positive tumors are enriched for cytokine-cytokine receptor interaction, cell cycles and IL-17 pathways. Unique DEGs in recurrent tumors were enriched for protein digestion and absorption, cAMP pathway. Shared DEGs were enriched for ECM-receptor interaction, Wnt signaling, PI3K-Akt, and MAPK signaling pathways.

**Supplemental Figure S4: Dynamic Changes in the Tumor Immune Landscape of the *MN1::PATZ1* Fusion-positive CNS Tumors.**

**A)** CIBERSORT suggests changes in immune cell population abundance among the primary and recurrent tumor, illustrating a shift in the immune microenvironment.

**B)** Immunohistochemistry staining of CD4 T cells in primary and recurrent samples. Scale bars = 100 μM.

**C)** UCSC Brain Tumor Map illustrating age distribution of cases.

**Supplemental Table S1: Clinical characteristics of pediatric patients.**

| Case ID | Diagnosis | Reported Driver Mutation | Age range (yrs)  Sex | Site | Treatment | Primary (P) /Recurrent (R) |
| --- | --- | --- | --- | --- | --- | --- |
| STN10126 | NET PATZ1 | *MN1::PATZ1* | 0-6  F | Left Frontal lobe | GTR | P |
| STN10156 | NET PATZ1 | *MN1::PATZ1* | 7-14  F | Left Frontal lobe | GTR | R |
| THR60_5285_S01 | Low-grade glioma | *MN1::PATZ1* | 0-6  M | Frontal lobe | Gross-Near total resection | P |
| THR_5286_S01 | High-grade glioma | *MN1::PATZ1* | 0-6  M | Thalamus | Partial resection, Radiation | R |
| STN10018 | PA | *PARK2* T240M  *SETD2* L263* | 7-14  F | Cerebellum | GTR | P |
| STN10047 | PA | None reported | 15-20  M | Cerebellum | GTR | P |
| STN10069 | PA | *KIAA1549::BRAF* | 0-6  F | Cerebellum | GTR | P |
| STN10086 | PA | *KIAA1549::BRAF* | 7-14  F | Posterior Fossa | GTR, Chemotherapy | R |
| STN10087 | PA | *KIAA1549::BRAF*  *CHEK2 p.T367f* | 7-14  M | Posterior Fossa | GTR | P |
| STN10088 | PA | *KIAA1549::BRAF* | 0-6  F | Posterior Fossa | GTR | P |
| STN10042 | Epilepsy | *None reported* | 0-6  F | Frontal lobe | Craniotomy | P |

**Supplemental Table S2: Summary of the UCSF 500 data to describe the fusion event on the exome level.**

|  | GRCh37/hg19 | GRCh38/hg38 |
| --- | --- | --- |
| UCSF 500-seq | Breakpoints in the intron 1 of *MN1* (chr22: 28,165,461) and exon 1 of *PATZ1* (chr22: 31,740,566). Annotated using RefSeq transcripts NM_002430.2 (Codons 1-1260) and NM_014323 (Codons 342-687), respectively. | Breakpoints in the intron 1 of *MN1* (chr22: 27,769,473) and exon 1 of *PATZ1* (chr22: 31,344,580) |

**Supplemental Table S3: Outliers for the primary, newly diagnosed *MN1::PATZ1* fusion tumor (STN10126). **Attached as separate file****

**Supplemental Table S4: Comparative Ingenuity Pathway Analyses of *MN1::PATZ1* fusion tumors (Group 1) with pilocytic astrocytoma (Group 2). **Attached as separate file****

**Supplemental Table S5: Adult type high-grade gliomas, including four glioblastomas, showing the topmost correlated RNA-seq samples to the primary *MN1::PATZ1* fusion-positive CNS tumor .**

| **Sample ID** | **Diagnosis (grade)** | **Histology** | **Age range (yrs)** | **Sex** |
| --- | --- | --- | --- | --- |
| TCGA-76-493101 | Glioblastoma (4) | Astrocytoma | 70-75 | F |
| TCGA-HT-A5RC-01 | Glioma (3) | Astrocytoma | 70-75 | F |
| TCGA-19-2619-01 | Glioblastoma (4) | Astrocytoma | 55-59 | F |
| TH40_2219_S01 | Supratentorial embryonal, NOS | PNET | 0-6 | M |
| TCGA-06-0158-01 | Glioblastoma (4) | Astrocytoma | 70-75 | M |
| TCGA-26-5139-01 | Glioblastoma (4) | Astrocytoma | 60-65 | F |

**Supplemental Table S6: Adult type intermediate-to-high-grade gliomas, showing the topmost correlated RNA-seq samples to the recurrent *MN1::PATZ1* fusion-positive CNS tumor.**

| **Sample ID** | **Diagnosis (grade)** | **Histology** | **Age (yrs)** | **Sex** |
| --- | --- | --- | --- | --- |
| TCGA-HT-A617-01 | Glioma (2) | Oligodendroglioma | 45-50 | M |
| TCGA-HT-7691-01 | Glioma (2) | Astrocytoma | 30-35 | F |
| TCGA-FG-A4MW-01 | Glioma (3) | Oligoastrocytoma | 60-65 | M |
| TCGA-P5-A72U-01 | Glioma (3) | Oligodendroglioma | 70-75 | F |
| TCGA-P5-A5F1-01 | Glioma (2) | Astrocytoma | 30-35 | M |
| TCGA-S9-A7IX-01 | Glioma (3) | Astrocytoma | 55-59 | M |

**Supplemental Table S7: Primary versus recurrent** ***MN1::PATZ1* fusion-positive CNS tumors edgeR results, relevant for the Volcano Blot in Figure 3A. **Attached as separate file****

**Supplemental Table S8: Whole transcriptomic data for primary and recurrent *MN1::PATZ1* fusion-positive CNS tumor (Stanford case) and five pilocytic astrocytoma (STN18, STN69, STN86, STN87, STN88), relevant for the Venn Diagram in Figure S3C**Attached as separate file****

**Supplemental Table S9: Summary of all published *MN1::PATZ1* cases until time of reporting**

| Study | Case # | Morphology | | Histopathology | | Radiology | Clinical Features | Treatment | FU (m) |
| --- | --- | --- | --- | --- | --- | --- | --- | --- | --- |
| Burel-Vandenbos^11^ | 1 | Malignant round cell  tumor-like | Rare perivascular pseudo-rosettes, hyaline stroma, infrequent microcystic growth pattern, focal endothelial cell proliferation, brisk mitotic activity | | Supratentorial right hemispheric, partly cystic & enhancing,  calciﬁcation | | 0-6 yrs F | Resection, 4 courses of CT, stem cell transplantation, RT (50 Gy) | 5 – no relapse |
| Rossi^12^ | 1 | High-grade glioma-like, oligodendroglial-like | Glia-like, round nuclei, hyalin, hypercellular foci (5-15%) with brisk mitoses, necrosis, pseudo-palisading, microvascular proliferation | | Parieto-occipital, lateral ventricle adjacent, prominent cystic component & enhancing, no edema, no calcification | | 0-6 yrs  F | GTR+CT | NED 18 |
| Rossi^12^ | 1 | Anaplastic, glioneuronal-like,  oligodendroglial-like | Glia-like, round nuclei, hyalin, hypercellular foci (5-15%) with brisk mitoses, necrosis, pseudo-palisading, microvascular proliferation | | Left parieto-occipital, lateral ventricle adjacent, prominent cystic component & enhancing, calcification, no edema | | 15-20 yrs  M | GTR+RT+CT | NED 64 |
| Rossi^12^ | 1 | Polymorphous, low-grade neuroepithelial tumor of the young-like, oligodendroglial-like | Glial-like, round nuclei, pleomorphic nuclei, low mitotic index, no anaplasia | | Frontal horn, lateral ventricle adjacent, cystic component & enhancing, calcification, no edema | | 15-20 yrs  F | GTR | NED12 |
| Tauziède-Espariat ^13^ | 1 | Glioneuronal-like | Polymorphous (spindle cells, pleomorphic, oligodendroglia-like), hyalin | | Right Lateral Ventricle | | 0-6 yrs F | GTR | In situ recurrence  PFS 139/OS 144 |
| Tauziède-Espariat ^13^ | 1 | Ependymoma-like | Polymorphous (pleomorphic, oligodendroglia-like), hyalin | | Right Parietal Lobe | | 0-6 yrs F | GTR+RT | No recurrence  PFS/OS 66 |
| Tauziède-Espariat **^13^** | 1 | Glioma-like | Polymorphous (pleomorphic, oligodendroglia-like), hyalin | | Pineal Lesion | | 0-6 yrs M | STR+CT | 1 in situ recurrence  PFS 4/OS 15 |
| Tauziède-Espariat ^13^ | 1 | Glioneuronal-like | Polymorphous (pleomorphic, oligodendroglia-like), hyalin | | Frontal lobe | | 45-50 yrs  M | GTR | No recurrence  PFS/OS 13 |
| Tauziède-Espariat ^13^ | 1 | Glioneuronal-like | Polymorphous (pleomorphic, oligodendroglia-like), hyalin | | Right Lateral Ventricle | | 15-20 yrs  F | STR | In situ recurrence  PFS 5/OS 20 |

CT, chemotherapy; GTR, gross total resection; NED, no evidence of disease; OS, overall survival; PFS, progression-free survival; RT, radiotherapy; STR, subtotal resection.
