## Supplementary material for "Glial Maturation and Immune Landscape Dynamics in *MN1::PATZ1* Fusion-Positive CNS Tumor Recurrence": supp tables and figure legends

**SUPPLEMENTAL FILE**

**Methods**

*Patient material and clinical information:* Appropriate institutional review board approved the project. Informed written consent was obtained from the patient’s parents prior to surgery. Patient specimen were given an ID number starting with STN, which are only known to the people within the research group. Two additional cases with *MN1::PATZ1* fusion-positive tumors were available from the Children’s Brain Tumor Network (CBTN) and were downloaded via the cpedsbioportal. Patient IDs were as follows: 1. PT_E3ADF4ZB, specimen BS_ZVZDDW2G (THR60_5285_S01), external patient identifier: C831849.

<https://pedcbioportal.kidsfirstdrc.org/patient/clinicalData?studyId=pbta_all&caseId=PT_E3ADF4ZB>. 2. PT_HZNZ635, specimen BS_49BQS7Z6 (THR60_5286_S01), external patient identifier: C801468**.**

<https://pedcbioportal.kidsfirstdrc.org/patient/clinicalData?studyId=pbta_all&caseId=PT_HNZNZ635>.

*Clinical information (Table S1):* A female pediatric patient (age range 0-6) presented after one week of fatigue, two days of headache, and vomiting and acute right hemiparesis on day of hospital admission. Neuroimaging revealed a large cystic tumor with ring enhancement. Gross-total resection was completed revealing a neuroepithelial tumor harboring an *MN1::PATZ1* fusion mutation, which was resected for a tumor recurrence 10 months later. Subsequent imaging demonstrated growth of the residual nodule located along the medial aspect of the resection cavity, with increased associated enhancement. There was also new enhancement in the anterior corpus callosum. The patient experienced a one-time seizure after the first surgery. As such, she was started on lacosamide, and future imaging studies were planned to monitor the aforementioned nodule, and to identify whether the enhancement in the anterior corpus callosum was related to seizure activity vs. residual tumor. Subsequent neuro-imaging indicated the enhancement in the anterior corpus callosum resolved, with residual T2/FLAIR brightness in that region, consistent with volume loss. However, the soft tissue nodule displayed growth and increased enhancement compared to more remote imaging, concerning for growth of residual tumor. This prompted the second surgical resection for the tumor recurrence. Pathology demonstrated neuroepithelial tumor with *MN1::PATZ1* fusion.

Overall, rapid post-operative recovery was noted. There were differences between the intra-and post-operative imaging, rendering comparison and interpretation of the current findings challenging. Thus, it was unclear whether the enhancing nodular lesion described above corresponded to residual disease vs. evolving post-operative changes, and watchful observation and shortened interval scans were therefore advised. Evidence of residual/progressive disease adjuvant irradiation of the resection cavity was noted to be considered.

*Targeted next-generation sequencing with the UCSF 500 Panel.*

Targeted Illumina sequencing was performed on paraffin embedded tissue using the UCSF 500 Cancer Gene Test (UCSF Pathology Laboratory), which generated 49 million paired-end 101 bp reads after cleaning and removal of duplicated reads. The *MN1::PATZ1* fusion had breakpoints in intron 1 of *MN1* annotated using RefSeq transcripts NM_002430(1) (breakpoint chr22:28165461) and in Exon 1 of PATZ1, annotated using RefSeq transcripts NM_014323 (breakpoint chr22:31740566). Genomic positions are given in reference to the human genome GRCh37/hg19 in UCSF 500 analysis pipeline and revealed an *MN1::PATZ1* fusion, with fusion junctions in intron 1 of *MN1* and exon 1 of *PATZ1* that caused an in-frame linkage of exon 1 of *MN1* (codons 1-1260) together with exons 1-5 of *PATZ1* (codons 342-687) annotated using RefSeq transcripts NM_002430.2 and NM_014323.

For validation, the paired-end reads were extracted from the alignment file from the original analysis, and 20 million 150bp PE raw reads were aligned to an alternative human reference genome GRCh38/hg38 using bwa-mem2. ^1^ More than 97% of these paired-end reads can uniquely and concordantly map to the reference genomes. Total mappability is about 99.6% against the GRCh38/hg38 reference genome (Table S2). This confirmation was achieved by 1.2 million (3%) of reads being chimeric or split reads, and being mate mapped to a different chromosome. The genome coordinates of the breakpoints are reference-dependent, but the fusion sequences and annotations from different reference genomes were consistent.

All testing was performed at UCSF Health Center for Clinical Genetics and Genomics using the UCSF 500 Cancer Gene Panel. The GTR Test ID is CGP-25048:T_ALT. Additional details can be found at [https://genomics.ucsf.edu/UCSF500](about:blank)

***Methylation profiling:***

The St. Jude’s pathology laboratory performs methylation profiling as a laboratory developed test in a CLIA/CAP certified environment using a custom algorithm  ^2^ which does not include the NET-PATZ tumor class.  Copy-number profiling from the Illumina DNA methylation arrays, the consume package ([http://bioconductor.org/packages/conumee/](https://urldefense.com/v3/__http:/bioconductor.org/packages/conumee/__;!!G92We9drHetJ8EofZw!aLGxnyXptsRntqGiIayUGpPWjyJnL1mO5FOjDmOUw2t2Y2K6pTzwbOPaCWV5QZCvh5JaBMOVsMN9qE_jDwfxIQ$)), was used as described (Figure S1C). ^3^

*RNA-based Analyses*

*Bulk RNA sequencing*

Resection samples were acquired from the operating room during surgery and were frozen within an hour after retrieval. Subsequently, these frozen resection samples of the primary tumor (STN10126) and its patient-matched recurrent tumor (STN10156) were submitted for bulk RNA-sequencing at Novogene. The RNA Integrity Number (RIN) was 6.1 and 6.7, for the primary and recurrent tumor, respectively. It passed the QC threshold and was subjected to paired-end sequencing. Total 61 and 62 million 150 bp paired-end sequencing reads were generated for STN10126 and STN10156, respectively. 70 million pair-end raw reads were aligned to the GRCh38/hg38 using hisat2 and STAR software. ^4,5^ Over 97% of these reads were successfully aligned, with unique and concordant mapping to the GRCh38/hg38 reference genome, achieving approximately 89% mappability.

*Tumor Mapping and Comparative Analysis of RNA expression (CARE)*

The UCSC TumorMap was used to visualize the Treehouse v11 polyA compendium, consisting of 12,747 pediatric and adult tumors, including 1,215 brain tumors (<https://doi.org/10.1038/srep25533>). ^6^ The method uses a spatial correlation analysis to plot clusters of tumors, which aids researchers in identifying similarities among groups of samples. Each hex in the *TumorMap* plot represents an individual sample. Placement of each hex is determined via pairwise similarity scores between samples causing samples with similar genomic profiles to be plotted near each other and form localized clusters on the *TumorMap*. Spearman rank correlation was used to determine the top six most correlated samples to the patient sample.

All RNAseq data were first uniformly processed using the Toil RNAseq pipeline version 3.2 developed by the UCSC Computational Genomics Lab (<https://doi.org/10.1038/nbt.3772>). We were unable to run the recurrent sample, because it had a low number of uniquely mapped reads and therefore did not meet the strict QC threshold for the Treehouse Analyses. Additionally, the two Children’s Brain Tumor Network Samples were excluded from this analysis as they were generated using a ribo-depleted library preparation method, potentially resulting in different values compared those generated by the polyA selection library preparation method, that was used for our pan-cancer and pan-disease compendia (Table S1). Gene-level transcript per million data was used to perform gene expression outlier analysis to identify significantly enriched genes in the patient’s tumor compared with 12,747 pediatric and adult tumors in the Tumor Compendium v11 Public PolyA. For pan-cancer analysis, we used the filtered set of 27,328 genes; for pan-disease analysis, we used the unfiltered set of 58,581 unique GENCODE Human Release 23 genes to make sure we did not miss genes whose expression is specific to certain tumor subtypes.

All samples included in the Treehouse Tumor Compendia and all samples with reported outlier expression results exceed our quality threshold of 10 million mapped, exonic, non-duplicate (MEND) reads. The threshold is based on the GEUVADIS Consortium minimum goal of 20 million reads ^7^ and our observation by the Treehouse Tumor Compendium group that the median fraction of MEND reads was 50% in a survey of 2,178 tumors from 48 cohorts. Based on these data, we at the Treehouse group use MEND reads as a quality control measure for RNA sequencing data. ^8^ Disease categories included in the compendium are choroid plexus carcinoma, embryonal tumor with multilayered rosettes, ependymoma, glioblastoma multiforme, glioma, gliomatosis cerebri, medulloblastoma, pineal parenchymal tumor, rosette forming glioneuronal tumor, and supratentorial embryonal tumor NOS brain tumors.

*Ingenuity Pathway Analyses (IPA)*

Pathway analysis of whole transcriptome datasets was conducted in Ingenuity Pathway Analysis (IPA) (v. 1-23-01, QIAGEN Inc., [https://digitalinsights.qiagen.com/IPA](https://nam02.safelinks.protection.outlook.com/?url=https%3A%2F%2Furldefense.com%2Fv3%2F__https%3A%2F%2Fdigitalinsights.qiagen.com%2FIPA__%3B!!NHLzug!IEyMnAPt0QarkHmSKDbF4S4rR6GR0o9QBpyE-PWa7A8_hbi6nNkO1pmSmKdJtjUHqomc_DvSApK7L4evBJJSEDs7OSs0Lg%24&data=05%7C02%7Cdaveam%40upmc.edu%7C538661f44d414715575008ddaf38b88d%7C8b3dd73e4e724679b19156da1588712b%7C0%7C0%7C638859380348863375%7CUnknown%7CTWFpbGZsb3d8eyJFbXB0eU1hcGkiOnRydWUsIlYiOiIwLjAuMDAwMCIsIlAiOiJXaW4zMiIsIkFOIjoiTWFpbCIsIldUIjoyfQ%3D%3D%7C0%7C%7C%7C&sdata=vPQqNMGbg9l1e3ducWGKw9n2YOfTfqeggGm1tZJrdrw%3D&reserved=0)). ^9^ Averaged count matrices for each gene were calculated from all samples of each Group 1, 2 and 3. The log of the ratio of the averaged gene counts to expression of the gene in a control sample (non-neoplastic brain obtained from epilepsy surgery, log[Avg count/control count]) was used as the “log fold change” for each gene detected. The log fold change relative to control of the top 8000 differentially expressed genes was used to generate IPA core analyses of expression profiles. Expression changes of key pathways were compared between each group by comparison analysis. Overall detected and predicted gene expression in canonical pathways is visualized by z-score for all significant comparisons (-logp>1.3). These pathways and molecular patterns are further grouped into overlapping networks with unique disease and biological functions.

*Differential Gene Expression (DGE), TIMER, Deconvolution-CIBERSORT*

Differential gene expression (DEG) analysis was performed using the ‘limma’ package to identify significantly dysregulated genes between experimental groups. Identified DEGs were subjected to functional enrichment analyses using Gene Ontology (GO), Kyoto Encyclopedia of Genes and Genomes (KEGG) with ‘clusterprofiler’ package to elucidate relevant biological processes, molecular functions, and signaling pathways. To further explore the functional relevance and pathway involvement of key DEGs, cnetplot from ‘enrichplot’ package was employed to visualize pathway–gene relationships.

*TIMER:* The immune cell infiltration levels of tumor samples were estimated using the Tumor Immune Estimation Resource (TIMER, http://timer.cistrome.org). The abundance of six major immune cell types between primary tumor and recurrent tumor (B cells, CD4⁺ T cells, CD8⁺ T cells, neutrophils, macrophages, and dendritic cells) was analyzed.

*CIBERSORT:* Immune cell infiltration and immune-related pathway activities were assessed through single-sample gene set enrichment analysis (ssGSEA, from ‘tidyverse’ and ‘GSVA’ package) and CIBERSORT, ^10^ enabling characterizations of immune landscape differences across groups. Data visualization was completed by utilizing the ‘ggplot2’ package. All analyses were conducted in R language within the R Studio environment.

*Venn Diagram*: Differential expression was computed with DESeq2 (median-of-ratios normalization; design ~ condition) for two contrasts: primary tumor vs pilocytic astrocytoma (PAs) control and recurrent chordoma vs PAs control. DEGs were defined at FDR < 0.05 and |log₂FC| ≥ 1 after filtering lowly expressed genes. Upregulated overlaps and condition-unique sets were derived in base R and visualized with ggvenn. For each gene set, KEGG pathway enrichment was performed using clusterProfiler (enrichKEGG; organism = “hsa”), mapping gene symbols to Entrez IDs via org.Hs.eg.db and top pathways are shown.

***Histologic processing, staining, immunohistochemistry, and evaluation***

Patient tissue was processed according to well-established histologic protocols including fixation in 10% neutral buffered formalin, paraffin embedding, sectioning (5-μm), and standard histochemical staining using hematoxylin and eosin (H&E), PAS, and reticulin. Comprehensive diagnostic evaluation included review by a board-certified pathologist specialized in anatomic pathology and neuropathology, as well as a senior fellow in neuropathology. Immunohistochemical stains were performed using antibodies against glial fibrillary acid protein (GFAP, clone GA-5, ready-to-use, BIOCARE, cat#PM065AA), and Synaptophysin (SYP, clone 27G2, ready-to-use, BIOCARE, cat#PM371AA), CD68 (clone KP1, ready-to-use, BIOCARE, cat#PMO33AA), CD163 (clone 10D6, ready-to-use, BIOCARE, cat#PM353AA), Desmin (clone D33, ready-to-use, BIOCARE, cat#PM036AA), CD4 (clone 4B12, ready-to-use, BIOCARE, cat#API3148AA), CD8 (clone C8/144B, ready-to-use, BIOCARE, cat#API3160AA), and proliferation marker KI67 (clone MIB-1, ready-to-use, BIOCARE, cat#API3156AA) with the 3,3'-diaminobenzidine (DAB) chromogen as a service by Histo-Tec Laboratory IncFor in-house immunohistochemical staining, OLIG2, clone 211F1.1, from Cell Marque, cat# 387M-16, was used at dilution 1:100 and staining was performed on Leica Bond III using Leica's ER2 solution for HIER and Bond Refine DAB Kit. PDL1, clone SP263, from Ventana, cat# 790-4905, was used as predilute and staining was performed on Ventana Ultra using Ventana's CC1 solution for HIER and Optiview DAB detection kit. IHC quality and quantification were independently verified by a neuropathologist.

Cryoprotected minced tissue from primary and recurrent tumors were embedded in Tissue-Tek O.C.T. Compound (Sakura Finetek USA, Inc., Torrance, CA) and stored at -80 °C until sectioned. 10 μm-thick sections were cut on a cryostat (Leica, Wetzlar, Germany) and placed on Fischerbrand Superfrost Plus slides (Menzel Glaser, VWR) and air dried for 1 h. Prepared slides were stored at -80°C. Frozen tissue slides containing primary and recurrent tumor sections were washed briefly with PBS and blocked with Blocking solution (0.3% Triton X-100, 10% normal donkey serum in PBS) for 1 h at room temperature (RT). Then the slides were incubated with primary antibodies at RT overnight in a humidified chamber. On the following day, the slides were washed briefly three times with PBS and incubated with secondary antibodies for 1 h at RT. Stained slides were washed three time with PBS and cover slipped with mounting solution (VectaShield H20002; Vector Laboratories, Newark, CA). The slides were imaged on a confocal microscope. The following primary antibodies were used: goat anti-Olig2 (Milipore: AB9610), 1:200; rabbit anti-NG2 (Sigma: AB5320), 1:200; Rat anti-MBP (Abcam: ab7349) 1:500. The secondary antibodies, all raised in donkey included: anti-rat Alexa 488, anti-goat Alexa 647 and anti-rabbit Alexa 594 (Jackson ImmunoResearch Labs, West Grove, PA), 1:200.

Whole slide confocal images were acquired at 10x objective. Images were imported into FIJI/ImageJ image analysis software for quantification of cellular density and fluorescence intensity of whole slide. All analyses were performed in a blinded fashion.
